# Unmet palliative care needs in England and Wales: population-based estimates and future projections (2025-2050)

**DOI:** 10.64898/2026.02.16.26345914

**Authors:** Therese Johansson, Katherine E Sleeman, Anne Finucance, Joanna M Davies, Lorna K Fraser, Irene J Higginson, Melanie Diggle, Fliss E M Murtagh, Anna E Bone

## Abstract

**Introduction:** With global populations ageing, demand for palliative care is increasing. Population-level assessments of unmet palliative care needs are essential for strategic planning, yet rigorous methods to estimate unmet needs are lacking. This study aimed to develop methods and estimate current and future population-level prevalence of unmet palliative care needs among adults in England and Wales.

**Methods:** Secondary analyses of data from a nationally representative post-bereavement survey in England and Wales in 2022 (n=1,194). Unmet needs in the survey sample were estimated using two methods: (1) reported unresolved symptoms and concerns using Integrated Palliative care Outcome Scale scores, cutoff ≥34/68; and (2) reported insufficient care provision from general practitioners. These methods were combined to further provide a conservative estimate (1 and 2) and a broad estimate (1 or 2). We examined associations with unmet needs using modified Poisson regression. Age-, gender- and nation-specific sample estimates were applied to mortality data for 2022 and projections from the Office for National Statistics to calculate population-level estimates and prevalence from 2025 to 2050.

**Results:** In 2022, 247,993 (46%) adult decedents in England and 17,209 (49%) in Wales had unmet palliative care needs using method 1; 244,612 (46%) and 15,280 (43%), respectively, using method 2. According to conversative and broad estimates, 32% and 61% could have unmet needs in England, and 29% and 62% in Wales. By 2050, prevalence of unmet needs are projected to rise by 21–26% in England and 14–19% in Wales depending on estimate used, with the largest absolute increase among those aged ≥85 years.

**Conclusions:** Unmet palliative care needs are high in England and Wales and projected to increase by 2050, regardless of method. We contrast methods based on unresolved symptoms and concerns or insufficient care provision, or both, to inform the planning and evaluating of equitable care.

**Key Messages:** *What is already known on this topic:*

- Although understanding population-level unmet palliative care needs is critical for effective service planning, robust and standardised methods to estimate these needs remain limited.

*What this study adds:*

- We use two methods to provide four estimates of population-level prevalence of unmet palliative care needs and discuss their strengths and limitations.
- Regardless of estimate, prevalence of unmet palliative care needs is high, ranging 32-61% in England and in 29-62% in Wales; the number of people with unmet needs is anticipated to increase by 21–26% in England and 14–19% in Wales by 2050.

*How this study might affect research, practice or policy:*

- This advancement in methods to estimate unmet palliative care needs can inform the development and evaluation of population-level strategies to improve end-of-life care.
- Our population-level estimates do not account for multiple long-term conditions which are rising and will likely increase the complexity of needs.
- There needs to be more investment in primary and community-based services to ensure high-quality symptom management and support for people and their families towards the end of life.

## Introduction

Worldwide, mortality patterns are changing as populations age and health-related suffering of non-communicable diseases continues to rise. Consequently, more people are likely to benefit from palliative care.^1-4^ Palliative care is a person-centred approach to care for people with life-limiting illness and their families, focusing on management of symptoms, alleviation of distress and improvement of quality of life. It can be delivered by specialist palliative care services, such as hospices, but is also commonly provided in primary and community care setting by generalists such as General Practitioners (GPs), community and district nurses, care home providers and hospital teams.^5^ Despite this, many people do not receive the care and support they need towards the end of life, which can have significant implications for their health, wellbeing, and comfort. Understanding and addressing palliative care needs and the extent to which these are met in a population is therefore essential for effective and equitable healthcare planning and service delivery. ^6 7^

A large body of work exists assessing palliative care needs in populations, often based on prevalence of diseases that are amenable to palliative, using administrative data such as death certificates.^8-10^ Some population-level palliative care needs assessments have also considered symptom burden^11^ and service utilisation,^12 13^ and others have included subjective measures of needs.^14^ However, very little research has attempted to quantify the extent or nature of unmet palliative care needs at the population level. Empirical evidence on how many people experience unmet palliative care needs, and characteristics of affected groups remains limited. In the literature, unmet needs are typically understood as the gap between the care required and the care received,^15^ though definitions and measurement approaches vary widely and few studies are based on representative samples or include populations with non-cancer diagnoses. ^15-17^

This study builds on earlier phases of the DUECare project, including a scoping review that identified three key domains for defining and measuring unmet needs: (1) symptoms and concerns; (2) access to services; and (3) sufficiency of service provision to address symptoms.^16^ Stakeholder workshops using nominal group technique further refined priorities for measurement, highlighting the importance of capturing both symptom-related and service-related dimensions of unmet palliative care needs.^18^ Through this work we developed a definition of unmet palliative care needs as occurring ‘when a person with life-limiting illness has symptoms, psychosocial concerns, or care requirements that are not adequately addressed through available services, with inability to access or receive person-centred care’.^18^

The aim of this study was to develop methods to estimate the prevalence of unmet palliative care needs among adults in England and Wales, examine associated socio-demographic and clinical factors, and project baseline and future population-level prevalence of adults with unmet palliative care needs to 2050.

## Materials and Methods

### Study design

Secondary analysis of a nationally representative mortality follow-back survey data sent to next of kin of people who died in 2022 in England and Wales,^19^ and of routinely available national death registry data and mortality projections.^20 21^ We report the study in line with Strengthening the Reporting of Observational Studies in Epidemiology (STROBE) recommendations.^22^

### Study data

#### Better End of Life survey data

As part of the Better End of Life study, a modified version of the QUALYCARE survey^23^ was conducted with adults who had registered the death of a family member six to ten months prior. A nationally representative sample of decedents were identified using stratified sampling applied to the ONS’ mortality data. Deaths were eligible if they had causes of death considered to be amenable to palliative care (ICD-10 codes sampled available in Supplement Table 1). The sampling process has been described in detail elsewhere.^19^ To ensure adequate power for regression analyses with 25 predictors, we applied a conservative rule of 20 observations per variable, indicating a minimum sample size of 550. Data were collected between May and November 2023 (n=1,194), for deaths occurring between August and December 2022. Details of the sampling frame composition and characteristics of decedents for whom responses were received are published on https://doi.org/10.6084/m9.figshare.31100068.

Proxy reports from bereaved family members offer a valuable lens for understanding unmet needs in the final weeks or months of life.^23 24^ While not without limitations, these accounts provide the best available population-based insights into symptom burden, care experiences, and perceived gaps in support. The Better End of Life survey collected data from bereaved family members on decedent socio-demographic and clinical characteristics (e.g., age, gender, financial circumstances, and cause of death). The survey also contained validated measures, including the Integrated Palliative care Outcome Scale (IPOS – proxy version), a 17-item measure assessing multidimensional symptoms and concerns across three sub-scales (*physical symptoms* (10 items), *emotional symptoms* (4 items), and *communication and practical issues* (3 items). ^25^ Each IPOS item relates to how a symptom or concern affected the person who died during their last week of life, rated using a 5-point Likert scale from 0 (‘not at all’) to 4 (‘overwhelmingly’ or ‘always’). Scores are summed up, with possible total scores ranging from 0 to 68. The survey also included questions about which health care services were received and whether enough help was received from these services.

#### National Mortality Registry Data

We extracted publicly availably mortality data for England and Wales from the Office for National Statistics (ONS)^20^ for 2022 (same year as deaths sampled in the Better End of Life survey), for adults aged 20 years and above (available age categories in the mortality data by cause of death precluded extraction for ages 18-19).

#### Official National mortality projections

Publicly available mortality projections for England and Wales for the years 2025 to 2050 were obtained from the ONS.^21^ We used the ‘principal projections’, which provides mid-year annual mortality projections by age and gender.

## Analysis

### Estimates from Better End of Life survey sample

To estimate the proportion of decedents experiencing unmet needs in the survey sample, we developed and applied two estimation methods informed by the approaches to measuring unmet palliative care needs identified in our scoping review and stakeholder consultation workshops: (1) unresolved symptoms and concerns; (2) insufficient care provision to address symptoms and concerns.^16 18^ The estimation methods were combined in different ways, generating four sample estimates, which are summarised in Box 1.

#### Estimate 1: Unresolved symptoms and concerns in the last week of life

Estimate 1 was based on reported unresolved symptoms and concerns, by using summed IPOS scores (scores of 0-4 across 17 items). A cut-off at the 50^*t*h^ percentile of the maximum possible total score range (≥34 out of 68) was applied to indicate unmet needs. A total score of >34 equates to being ‘moderately’ affected by all items or ‘overwhelmingly/always’ affected for at least nine items. Observations with eight or more IPOS items missing were excluded from the analytic sample and examined in a sensitivity analysis. For observations with one to seven IPOS items missing, we used mean scores from the non-missing items to impute missing values.

#### Estimate 2: Insufficient care provision in the last 3 months of life

For estimate 2, unmet palliative care needs were indicated by reported *insufficient care provision*. In this study, we focus on primary palliative care, which is palliative care provided by care professionals working in primary and community-based care services.^26 27^ Primary palliative care should be available to everyone with advanced illness, and involves palliative care delivery integrated with chronic disease management.^28^ We did not focus on specialist palliative care provision, because it was not possible in our dataset to distinguish between those who did not need specialist palliative care and those who needed it but did not receive it. Estimate 2 was based on the survey question *“In the last three months before they died, overall, do you feel they got as much help as needed from GPs?”*. Responses of ‘no’, ‘sometimes’, or ‘did not receive care’ were coded as having unmet needs (i.e., not receiving enough help or not receiving care at all); ‘yes, most of the time’ was coded as having needs met (i.e., receiving enough help). Given that the survey sampled causes of deaths considered amenable to palliative care (i.e., expected deaths), we assumed decedents would have required at least some primary care in their final months of life.

#### Estimate 3: Conservative estimate

We calculated a conservative estimate based on the proportion of decedents who had unmet palliative care needs identified by both estimation methods 1 and 2 (i.e., those with a high burden of symptoms and concerns who also did not receive enough, or any, help from GPs).

#### Estimate 4: Broad estimate

A broad estimate was calculated based on decedents identified by either estimation method 1 or 2 (i.e., those with either a high symptom burden or those who did not receive enough help from GPs).

For each sample estimate, we calculated the 95% confidence intervals based on proportions using the Wilson score method.^29^ For each estimate we excluded observations with information about decedent gender and/or age missing.

#### Regression analysis

We report frequences and proportions for categorical variables and means with standard deviations for continuous variables. Associations between unmet need and socio-demographic or clinical characteristics were examined using modified Poisson regression with robust error variance.^30^ Separate multivariable models were fitted for binary outcomes (unmet needs vs no unmet needs) derived using the main two estimation methods, adjusting for gender, age, ethnicity, financial circumstances, nation, cause of death, and comorbidities. We report unadjusted and adjusted prevalence ratios (PR) with 95% confidence intervals. Analyses were performed in Stata 18.0.^31^

##### Box 1.

*Overview of how estimates of unmet palliative care needs are derived in this study*.

1. **Unresolved symptoms and concerns** An individual is identified as having unmet palliative care needs if they have an IPOS total score ≥34 (≥50% of maximum possible score).
2. **Insufficient care provision** An individual is identified as having unmet palliative care needs if it is reported that they did not receive enough, or any, help from GPs.
3. **Conservative estimate (1 and 2 combined)** An individual is identified as having unmet palliative care needs if they have an IPOS total score ≥34 **and** did not receive enough, or any, help from GPs.
4. **Broad estimate (either 1 or 2)** An individual is identified as having unmet palliative care needs if they either have an IPOS total score ≥34 **or** did not receive enough, or any, help from GPs.

### Population-level estimates

To estimate unmet palliative care needs among all adult deaths (age >20) in England and Wales, we first identified the population considered ‘at risk’ in the ONS mortality data for 2022^21^ based on the main expected causes of death considered amenable to palliative care that were sampled in the Better End of Life survey (Supplement table 1). We assumed that those dying from other causes of death (including sudden deaths and accidents) did not have unmet palliative care needs. We grouped these deaths by gender (male and female) and age (20-64, 65-84, ≥85 years) for each nation separately. For each of the four sample estimates, we then applied age-, gender- and nation-specific proportions to the ‘at risk’ deaths in England and Wales to derive population-level estimates of unmet palliative care needs. We report these as numbers and proportions of decedents with unmet palliative care needs out of all adult deaths in each nation. In addition to the point estimates, we derived upper and lower population-level estimates by applying the upper and lower limits of the 95% confidence intervals of each sample estimate to the population data.

### Future projections

We estimated future trends in unmet palliative care needs for adults in England and Wales over a 25-year period (2025–2050), using ONS national mortality projections.^21^ Age-, gender- and nation-specific population-level estimates were applied to projected mortality data for all adult deaths, using age groups 20-64, 65-84, ≥85 years. We assumed that the distribution of causes of death and the proportion of unmet palliative care needs remained constant throughout the projection period.

### Patient and public involvement

This study is part of the Marie Curie-funded DUECare project (MCCRP-23-02), which has had input from our patient and public involvement (PPI) group throughout. Our operationalisation of methods to estimate unmet palliative care needs was informed by discussions with our PPI representatives and built on findings from stakeholder workshops conducted in an earlier phase of the project.^18^

### Ethics

The study was conducted in accordance with the ethical standards set out in the Declaration of Helsinki and its amendments.^32^ Ethical approval for the Better End of Life survey was received from King’s College London Research Ethics Committee (HR/DP-21/22-24690). The analysis of routinely collected aggregated and anonymised data that are publicly available does not require ethical approval.

## Results

### Sample estimates

The mean age of decedents in the Better End of Life survey sample (n=1,194) was 81.5 years; 51.9% were female, 97.4% were White. There was a similar distribution of decedents across England (n=619, 51.8%) and Wales (n=575, 48.2%). The most common single cause of death was cancer (28.5%), followed by heart disease (15.3%) and dementia (15.3%). Full sample characteristics are presented in Table 1. Twenty-one observations missing information about decedent gender and/or age were removed from the sample estimations, along with observations with missing outcome for either method 1 (n=42) or method 2 (n=46). Overall, IPOS data was imputed for 316 observations with one to seven missing IPOS items.

**Table 1.**
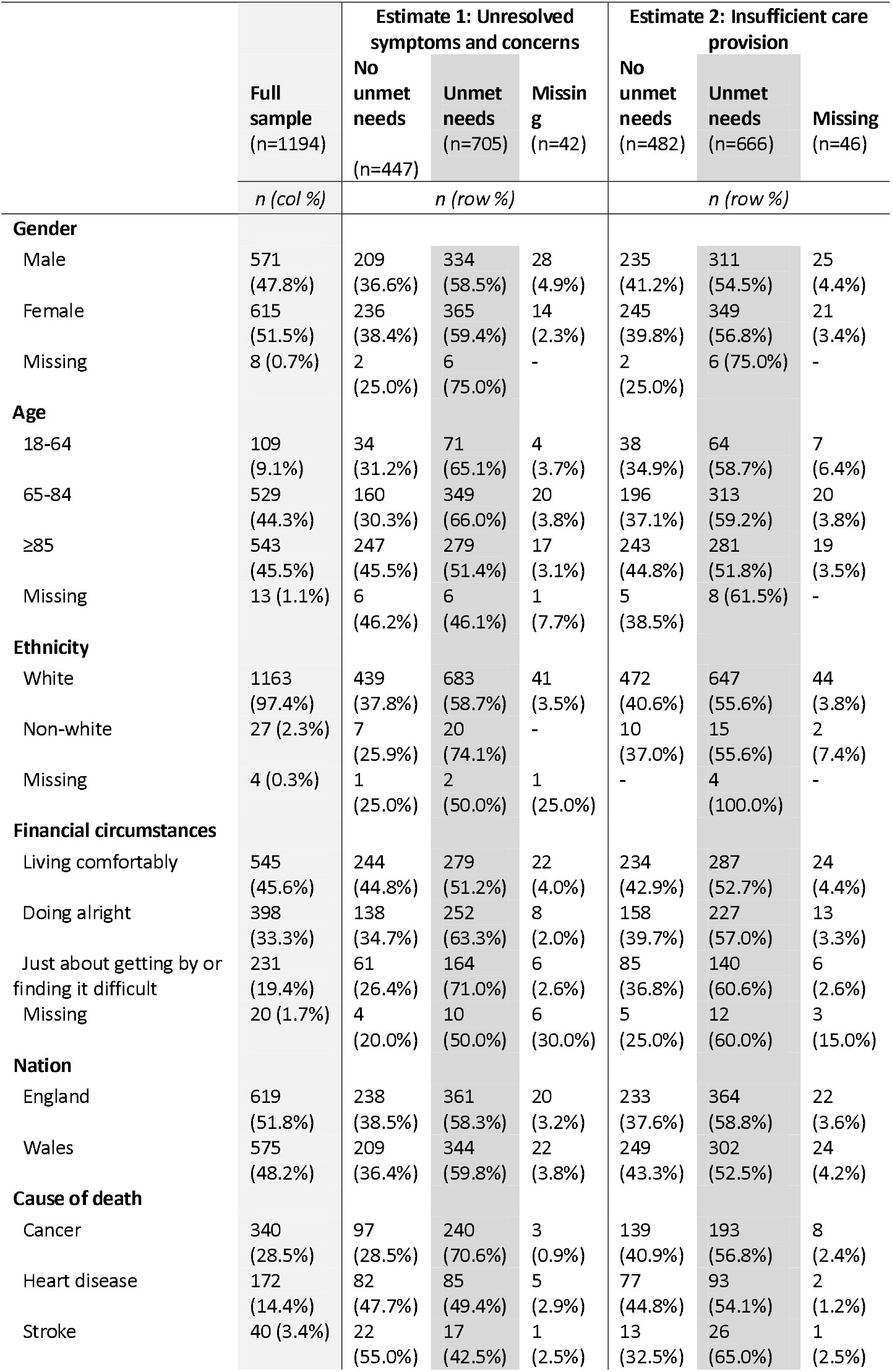
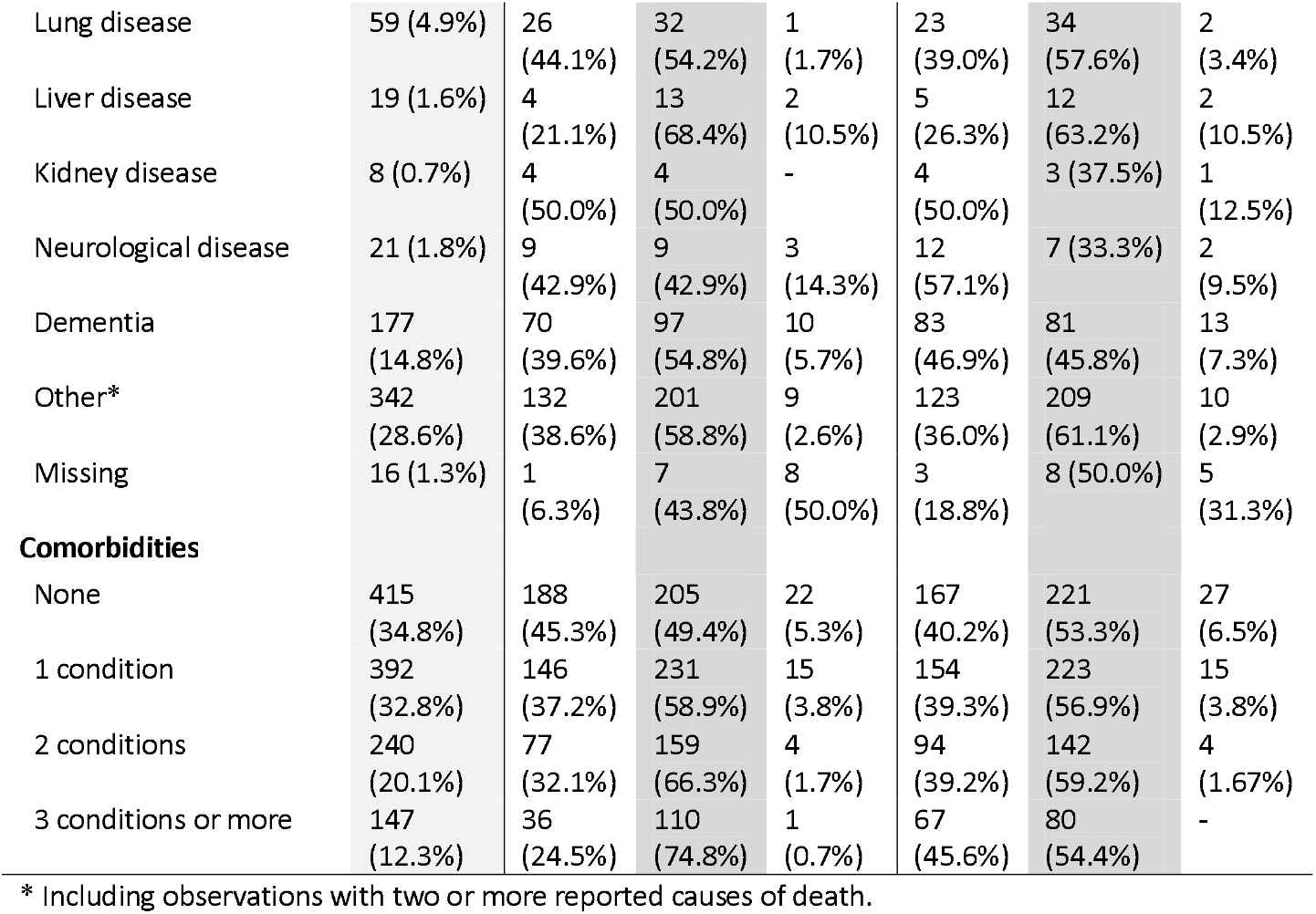
Sociodemographic and clinical characteristics of decedents in the Better End of Life survey sample (n=1,194).

|  |  | Estimate 1: Unresolved symptoms and concerns |  |  | Estimate 2: Insufficient care provision |  |  |
| --- | --- | --- | --- | --- | --- | --- | --- |
|  | Full sample<br>(n=1194) | No unmet needs<br>(n=447) | Unmet needs<br>(n=705) | Missing<br>(n=42) | No unmet needs<br>(n=482) | Unmet needs<br>(n=666) | Missing<br>(n=46) |
|  | *n (col %)* | *n (row %)* |  |  | *n (row %)* |  |  |
| Gender |  |  |  |  |  |  |  |
| Male | 571<br>(47.8%) | 209<br>(36.6%) | 334<br>(58.5%) | 28<br>(4.9%) | 235<br>(41.2%) | 311<br>(54.5%) | 25<br>(4.4%) |
| Female | 615<br>(51.5%) | 236<br>(38.4%) | 365<br>(59.4%) | 14<br>(2.3%) | 245<br>(39.8%) | 349<br>(56.8%) | 21<br>(3.4%) |
| Missing | 8 (0.7%) | 2<br>(25.0%) | 6<br>(75.0%) | - | 2<br>(25.0%) | 6 (75.0%) | - |
| Age |  |  |  |  |  |  |  |
| 18-64 | 109<br>(9.1%) | 34<br>(31.2%) | 71<br>(65.1%) | 4<br>(3.7%) | 38<br>(34.9%) | 64<br>(58.7%) | 7<br>(6.4%) |
| 65-84 | 529<br>(44.3%) | 160<br>(30.3%) | 349<br>(66.0%) | 20<br>(3.8%) | 196<br>(37.1%) | 313<br>(59.2%) | 20<br>(3.8%) |
| ≥85 | 543<br>(45.5%) | 247<br>(45.5%) | 279<br>(51.4%) | 17<br>(3.1%) | 243<br>(44.8%) | 281<br>(51.8%) | 19<br>(3.5%) |
| Missing | 13 (1.1%) | 6<br>(46.2%) | 6<br>(46.1%) | 1<br>(7.7%) | 5<br>(38.5%) | 8 (61.5%) | - |
| Ethnicity |  |  |  |  |  |  |  |
| White | 1163<br>(97.4%) | 439<br>(37.8%) | 683<br>(58.7%) | 41<br>(3.5%) | 472<br>(40.6%) | 647<br>(55.6%) | 44<br>(3.8%) |
| Non-white | 27 (2.3%) | 7<br>(25.9%) | 20<br>(74.1%) | - | 10<br>(37.0%) | 15<br>(55.6%) | 2<br>(7.4%) |
| Missing | 4 (0.3%) | 1<br>(25.0%) | 2<br>(50.0%) | 1<br>(25.0%) | - | 4<br>(100.0%) | - |
| Financial circumstances |  |  |  |  |  |  |  |
| Living comfortably | 545<br>(45.6%) | 244<br>(44.8%) | 279<br>(51.2%) | 22<br>(4.0%) | 234<br>(42.9%) | 287<br>(52.7%) | 24<br>(4.4%) |
| Doing alright | 398<br>(33.3%) | 138<br>(34.7%) | 252<br>(63.3%) | 8<br>(2.0%) | 158<br>(39.7%) | 227<br>(57.0%) | 13<br>(3.3%) |
| Just about getting by or finding it difficult | 231<br>(19.4%) | 61<br>(26.4%) | 164<br>(71.0%) | 6<br>(2.6%) | 85<br>(36.8%) | 140<br>(60.6%) | 6<br>(2.6%) |
| Missing | 20 (1.7%) | 4<br>(20.0%) | 10<br>(50.0%) | 6<br>(30.0%) | 5<br>(25.0%) | 12<br>(60.0%) | 3<br>(15.0%) |
| Nation |  |  |  |  |  |  |  |
| England | 619<br>(51.8%) | 238<br>(38.5%) | 361<br>(58.3%) | 20<br>(3.2%) | 233<br>(37.6%) | 364<br>(58.8%) | 22<br>(3.6%) |
| Wales | 575<br>(48.2%) | 209<br>(36.4%) | 344<br>(59.8%) | 22<br>(3.8%) | 249<br>(43.3%) | 302<br>(52.5%) | 24<br>(4.2%) |
| Cause of death |  |  |  |  |  |  |  |
| Cancer | 340<br>(28.5%) | 97<br>(28.5%) | 240<br>(70.6%) | 3<br>(0.9%) | 139<br>(40.9%) | 193<br>(56.8%) | 8<br>(2.4%) |
| Heart disease | 172<br>(14.4%) | 82<br>(47.7%) | 85<br>(49.4%) | 5<br>(2.9%) | 77<br>(44.8%) | 93<br>(54.1%) | 2<br>(1.2%) |
| Stroke | 40 (3.4%) | 22<br>(55.0%) | 17<br>(42.5%) | 1<br>(2.5%) | 13<br>(32.5%) | 26<br>(65.0%) | 1<br>(2.5%) |
| Lung disease | 59 (4.9%) | 26<br>(44.1%) | 32<br>(54.2%) | 1<br>(1.7%) | 23<br>(39.0%) | 34<br>(57.6%) | 2<br>(3.4%) |
| Liver disease | 19 (1.6%) | 4<br>(21.1%) | 13<br>(68.4%) | 2<br>(10.5%) | 5<br>(26.3%) | 12<br>(63.2%) | 2<br>(10.5%) |
| Kidney disease | 8 (0.7%) | 4<br>(50.0%) | 4<br>(50.0%) | - | 4<br>(50.0%) | 3 (37.5%) | 1<br>(12.5%) |
| Neurological disease | 21 (1.8%) | 9<br>(42.9%) | 9<br>(42.9%) | 3<br>(14.3%) | 12<br>(57.1%) | 7 (33.3%) | 2<br>(9.5%) |
| Dementia | 177<br>(14.8%) | 70<br>(39.6%) | 97<br>(54.8%) | 10<br>(5.7%) | 83<br>(46.9%) | 81<br>(45.8%) | 13<br>(7.3%) |
| Other* | 342<br>(28.6%) | 132<br>(38.6%) | 201<br>(58.8%) | 9<br>(2.6%) | 123<br>(36.0%) | 209<br>(61.1%) | 10<br>(2.9%) |
| Missing | 16 (1.3%) | 1<br>(6.3%) | 7<br>(43.8%) | 8<br>(50.0%) | 3<br>(18.8%) | 8 (50.0%) | 5<br>(31.3%) |
| **Comorbidities** |  |  |  |  |  |  |  |
| None | 415<br>(34.8%) | 188<br>(45.3%) | 205<br>(49.4%) | 22<br>(5.3%) | 167<br>(40.2%) | 221<br>(53.3%) | 27<br>(6.5%) |
| 1 condition | 392<br>(32.8%) | 146<br>(37.2%) | 231<br>(58.9%) | 15<br>(3.8%) | 154<br>(39.3%) | 223<br>(56.9%) | 15<br>(3.8%) |
| 2 conditions | 240<br>(20.1%) | 77<br>(32.1%) | 159<br>(66.3%) | 4<br>(1.7%) | 94<br>(39.2%) | 142<br>(59.2%) | 4<br>(1.67%) |
| 3 conditions or more | 147<br>(12.3%) | 36<br>(24.5%) | 110<br>(74.8%) | 1<br>(0.7%) | 67<br>(45.6%) | 80<br>(54.4%) | - |

\* Including observations with two or more reported causes of death.

For estimation method 1 (*unresolved symptoms and concerns*), the mean total IPOS score was 36.1 (SD 12.2, ranging 0 to 66.9). Overall, 696 of 1135 (61.3%) decedents had a score ≥34, indicating unmet palliative care needs (see Supplement file 2 for details about score distribution, IPOS data completeness, and descriptives of the excluded group with ≥8 missing IPOS items).

Using estimation method 2 (*insufficient care provision*), 655 of 1130 (58.0%) decedents were reported to not have received enough help or no help at all from GPs in the last three months of life (response distribution provided in Supplement Table 3).

After combining estimation methods 1 and 2 (see Supplement Table 4), 429 of 1104 decedents (38.9%) were identified as having have unmet palliative care needs using the conservative estimate (both method 1 and 2). The broad estimate (either method 1 or 2) identified that 886 of 1104 (80.3%) had unmet palliative care needs. Prevalence of unmet palliative care needs by decedent characteristics are reported for estimate 1 and 2 in Table 1 (Supplement Table 5 shows prevalence by decedent characteristics using estimates 3 and 4). Age-, gender- and nation-specific proportions for each sample estimate are provided in Supplement Table 6.

### Associations between decedent characteristics and unmet palliative care needs

#### Estimation method 1: Unresolved symptoms and concerns

In the unadjusted model, there was higher prevalence of unmet needs among decedents aged 18-64 and 65–84 compared to those aged 85 years or older. This association slightly attenuated in the adjusted model and only remained statistically significant for those aged 65–84. A gradient between unmet palliative care needs and financial circumstance was observed in the unadjusted model, with higher prevalence among decedents who were *“just about getting by or finding it difficult”* or *“doing alright”* compared to those *“living comfortably”*. This gradient remained significant in the adjusted model. Prevalence of unmet needs was higher among decedents who died from cancer compared to those who died from other conditions in both the unadjusted model and adjusted model. A gradient was also observed between unmet needs and comorbidities, with prevalence of unmet needs increasing with higher numbers of comorbidities. This association was slightly strengthened after adjustment.

#### Estimation method 2: Insufficient care provision

In the unadjusted model, decedents aged 65–84 had higher risk of unmet palliative care needs compared with those aged 85 years or older, and this association slightly strengthened after adjustment. Compared to those *“living comfortably”*, unmet needs were more prevalent among decedents who were *“just about getting by or finding it difficult”*. However, this association was attenuated in the adjusted model. Prevalence of unmet needs was lower among decedents in Wales compared to those in England, though this association attenuated and was no longer statistically significant in the adjusted model. Compared to those who died from other conditions, those who died from dementia had lower prevalence of unmet needs, and this association remained significant after adjustment.

The PRs for the decedent characteristics in each adjusted model are illustrated in Figure 1. Results from both unadjusted and adjusted models are presented in Supplementary table 7.

**Figure 1.**
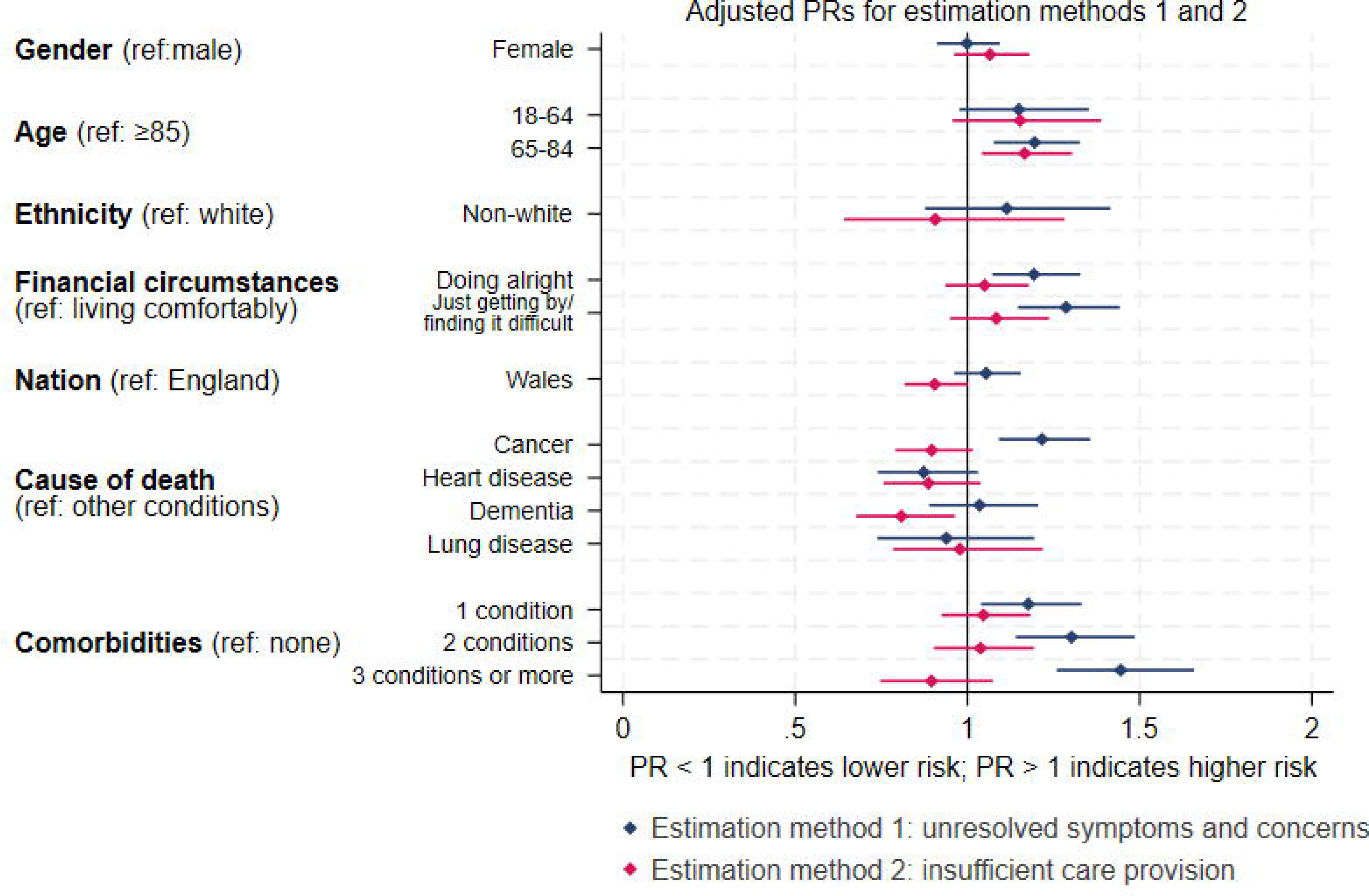
Forest plot of adjusted prevalence ratios (PR) with 95% confidence intervals for associations between decedent characteristics and unmet palliative care needs using estimation methods 1 and 2. Notes: Prevalence ratios adjusted for all covariates in the figure.

### Population-level estimates

In 2022, there were 536,311 adult deaths (age 20+) in England and 35,511 in Wales. Of those, 404,995 (75.5%) and 26,996 (76.0%) decedents, respectively, died from one of the main causes of death considered amenable to palliative care (ICD-10 codes listed in Supplement table 1). The four population-level point estimates of unmet palliative care needs, with upper and lower estimates based on the 95% confidence intervals of sample estimates, are reported separately for England and Wales in Table 2. Age-, gender- and nation-specific population-level estimates of unmet needs are reported in Supplement table 8.

**Table 2.** Estimated numbers and proportions of adult decedents (≥20) in England and Wales with unmet palliative care needs in 2022 using the four estimates.

| Estimate | England<br>(N=536,311) |  | Wales<br>(N=35,511) |  |
| --- | --- | --- | --- | --- |
|  | Estimated *n*<br>(%) | Lower–Upper<br>estimate* (%) | Estimated<br>*n* (%) | Lower–Upper<br>estimate* (%) |
| 1. Symptoms and concerns | 247,993<br>(46.2) | 210,506–285,479<br>(39.3–53.2) | 17,209<br>(48.5) | 14,576–19,841<br>(41.0–55.9) |
| 2. Insufficient care provision | 244,612<br>(45.6) | 205,812–283,412<br>(38.4–52.8) | 15,280<br>(43.0) | 12,575–17,986<br>(35.4–50.6) |
| 3. Conservative estimate<br>(1 and 2 combined) | 169,537<br>(31.6) | 129,959–209,116<br>(24.2–39.0) | 10,246<br>(28.9) | 7,536–12,956<br>(21.2–36.5) |
| 4. Broad estimate (either | 325,113 | 293,657–356,568 | 22,109 | 20,063–24,156 |
| 1 or 2) | (60.6) | (54.8–66.5) | (62.3) | (56.5–68.0) |

\* Lower and upper estimates for population prevalence derived from applying the upper and lower 95% confidence interval limits for each sample estimate to the population data.

### Future projections (2025-2050)

Applying age- and gender-specific estimates to national mortality projections indicates that the number of adults in England and Wales with unmet palliative care needs will rise markedly over the next 25 years. By 2050, the annual number of adults with unmet palliative care needs is projected to increase by 21–26% in England and by 14–19% in Wales compared with 2025, depending on the estimation method used (see Figure 2). Depending on the estimate used, it is projected that the number of people with unmet palliative care needs will increase by 39,000–78,700 in England and by 1,400–4,300 in Wales between 2025 and 2050. The greatest absolute increase in prevalence is expected among adults aged 85 years or older (see Supplement table 9).

**Figure 2.**
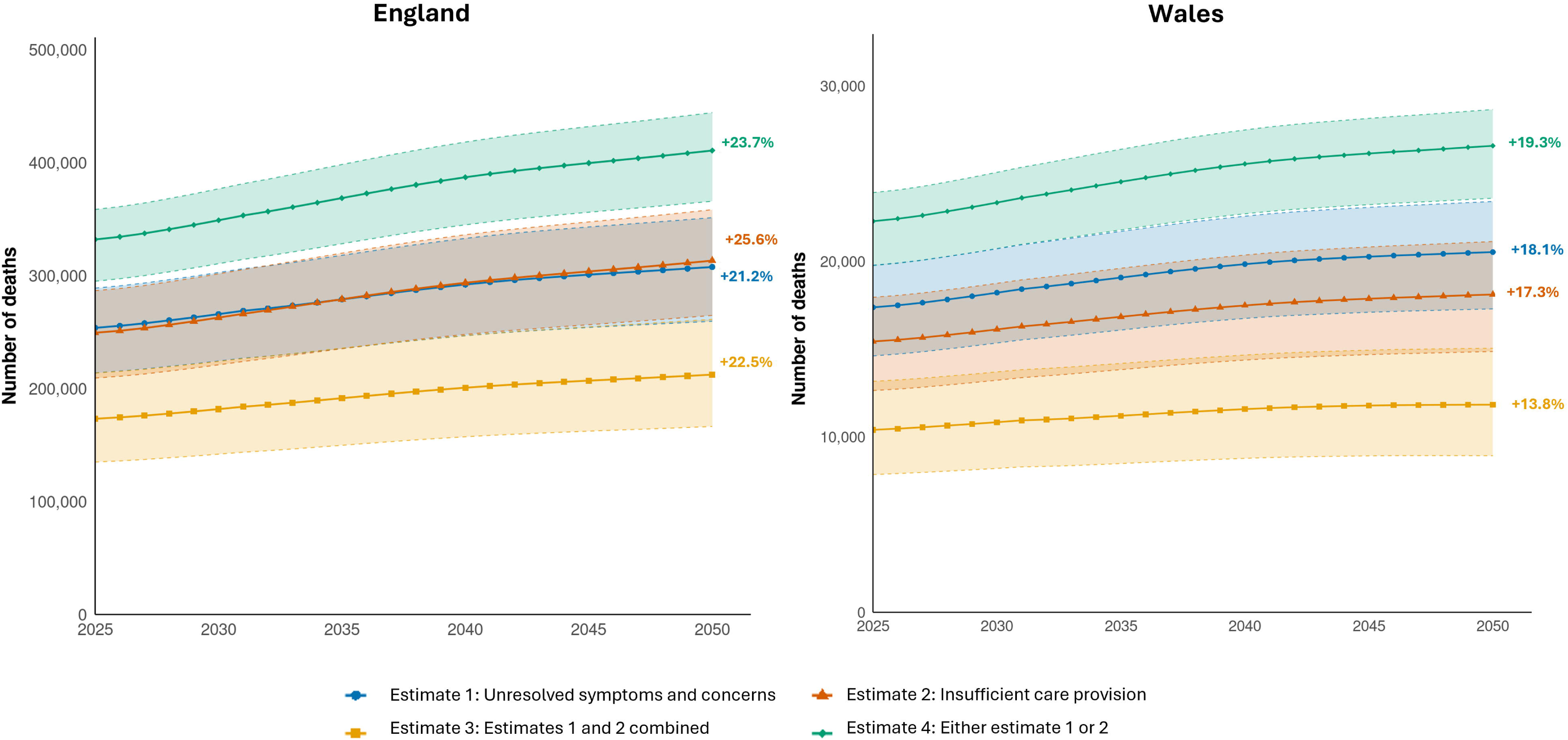
Projected prevalence of unmet palliative care needs among adult decedents (≥20) in England and Wales (2025–2050), based on levels in 2022 (assuming no major policy or service changes). Notes: The shaded area for each plot illustrates upper and lower population-level estimates applied to mortality projections

## Discussion

To our knowledge, this is the first study to develop robust empirical estimates of unmet palliative care needs and apply these to all adult decedents at a nation-level. We present two estimation methods and four estimates of unmet palliative care needs, developed from existing literature and with stakeholder engagement, accounting for both objective and subjective aspects of the construct. We show that prevalence of unmet palliative care needs is high, irrespective of estimate used: between a third and two-thirds of all deaths in England and Wales in 2022 are estimated to have unmet palliative care needs. The risk of having unmet palliative care needs was higher among those aged 65-84, people in poorer financial circumstances, and those who had a higher number of comorbidities. Future unmet palliative care needs are projected to increase markedly, by 21–26% in England and 14–19% in Wales by 2050, with the largest absolute rise among adults aged 85 years or older.

Our findings suggest that even in the UK, which has a well-developed system of palliative care, provision for people approaching the end of life is falling short. Workforce planning and resource allocation must account for the projected rise in unmet palliative care needs, especially among the oldest age groups, where mortality will increase most rapidly, and in primary care services. This will be important if the ambitions of ‘ageing in place’ and shifting from acute to community-based care outlined in health care policy recommendations in the UK^33 34^ and internationally^35-37^ are to be achieved. GPs and community nurses already provide a large part of the care dying people receive in community settings,^38^ and patients and families often expect that their GP will be involved in provision of end-of-life care.^39^ The prevalence of unmet needs using estimation method 2, which concerns primary palliative care (specifically GP provision) was high in this study (45.6% in England and 43.0% in Wales), and the ongoing shift to more community-based care will increase pressures on primary and community-based care services further. A previous comparative study of 16 countries also identified private homes and nursing homes as the settings where need for palliative care provision is the highest.^40^ Integration of palliative care into primary care is also highlighted by the World Health Organization as a crucial step to improve access to palliative and end-of-life care globally.^41^

Our study makes several important contributions to the evidence base on unmet palliative care needs. We address an evidence gap concerning the lack of operational clarity in measuring unmet palliative care needs identified in recent reviews.^15 16 42^ We advance the field by developing two estimation methods to generate four clearly operationalised estimates to quantify unmet palliative care needs, and demonstrate how these compare within one nationally representative sample of decedents who died from causes of death amenable to palliative care. These methods for estimation offer potential for use in other nations with comparable health systems.

While no estimation method is perfect, person-centred outcome measures completed by patients are arguably best current practice for assessing unmet palliative care needs. Service access measures are often used as an indicator of unmet palliative care needs at the population level^43^ but are limited as they do not capture whether the services accessed were effective in addressing patients’ symptoms and concerns. For example, in 2011 it was estimated that of all annual deaths in England with palliative care needs, 20-25% were not receiving specialist palliative care and thus had unmet needs.^17^ These estimations, however, did not regard of subjective experiences of unmet palliative care needs. Routine national collection of validated person-centred outcome measures that identify the nature of a patient’s symptoms and concerns and the extent to which care has resolved or improved them may therefore provide the most comprehensive measurement of unmet palliative care needs (aligning with the conservative estimate in this study). Such measures would provide important opportunity to monitor care quality and improve service delivery.^44 45^ Despite the availability of numerous validated tools designed to assess unmet palliative care needs,^42 46^ their implementation into clinical practice remains limited. However, development of a prototype national Palliative Care Outcomes Registry,^47^ together with evidence from other countries^48 49^ suggest that routine use of person-centred outcome measures is feasible. Inclusion of self-report measures of unmet palliative care needs in nationally representative surveys is another possible means of assessment.^50 51^

Several socio-demographic and clinical factors were associated with the prevalence of unmet palliative care needs across both estimation method 1 (*unresolved symptoms and concerns*) and 2 (*insufficient care provision*). Patterns were broadly consistent between methods, except for comorbidities where method 1 showed a clear gradient of higher prevalence of unmet needs and with higher number of comorbidities, whereas method 2 did not. This divergence likely reflects limitations with capturing the relationship using method 2 rather than actual lower prevalence among people with multiple conditions, given previous research findings of high levels of healthcare consumption and unmet needs for primary care for this group.^52-54^ A gradient was also observed between unmet needs and financial hardship for both methods, though only statistically significant for method 1, which aligns with existing evidence that people with lower socioeconomic position experience poorer healthcare outcomes and greater symptom burden towards the end of life.^55 56^

For both estimation methods, unmet needs were more prevalent among decedents aged 65-84 than those aged 85 or older. This could in part be due to reduced insight about GP care provision among survey respondents reporting for relatives in the oldest age group, many of whom lived in care homes. This explanation might also relate to the lower prevalence of unmet needs among those who died from dementia using method 2, which diverges from research identifying barriers to care access for people with dementia^57^. No association between ethnicity and the risk of unmet palliative care needs was observed in this study, likely due to the small number of non-White British decedents in the Better End of Life survey sample and the use of binary ethnicity categories. The proportion of white decedents in our sample reflects national demographic patterns for older populations in England and Wales,^58^ however the aggregation of non-white ethnicities is likely to mask important differences between ethnic minority groups. In future surveys, attempts to oversample in areas of high ethnic diversity could help to provide more data to explore differences in unmet need by ethnicity.

### Strengths and limitations

A key strength of this study is the application of multiple approaches to estimate unmet palliative care needs. Our two estimation methods operationalise different aspects of unmet palliative care needs, adopting a needs-based rather than diagnosis-based perspective that is more closely aligned with the palliative care approach. The estimation methods were based on existing measurement approaches identified in our scoping review and informed by stakeholder-prioritised elements of unmet needs. We derived our sample estimates from the Better End of Life survey, the first nationally representative post-bereavement survey in England and Wales since VOICES in 2015.^59^

Some limitations to this study should be noted. First, the Better End of Life survey was not designed to directly measure *unmet* palliative care needs. The sample estimates applied here should be tested in other studies, to compare findings and address limitations identified for this study. Survey data comprise proxy-reports by bereaved family members; although non-family respondents are included, decedents without family are less well represented in these estimates. Furthermore, respondents who were not closely involved in their relative’s care might have less insight into their needs and whether these were met or not. IPOS data on unresolved symptoms and concerns used in estimation method 1 relate to the last week of life; while this does not capture the extent to which needs were addressed earlier in the illness trajectory, it remains a proxy for good prior managements. Real-time recording of symptoms and concerns over time would allow for more comprehensive understanding of individual needs for palliative and end-of-life care and the extent to which these are met. For estimation method 2, we only focused on primary palliative care (i.e., receiving enough help from GPs in the last 3 months of life). A method for estimating unmet needs for specialist palliative care services specifically is an important area for future research.

There are also limitations for the population-level estimates. We only consider unmet palliative care needs at the end of life among people who died; unmet palliative care needs earlier in the illness trajectory are not reflected in these estimates. We may be missing unmet needs among other causes of death that are non-sudden but not included in the list of ICD-10 codes. It should be noted that for the projections we assumed that the distribution of causes of death would remain constant over time and did not account for rising of prevalence of major disease groups such as cancer and dementia. Projections also do not account for multiple-long term conditions, which is common, especially in older age groups,^60 61^ and will continue to grow, likely increasing the complexity of needs. Importantly, we assumed in the projections that service delivery will remain stable. In the UK, hospices are increasingly struggling with funding, with a rise in closures, which will likely have knock-on effects on care access. Taken together, these considerations mean that population-level estimates and future projections of unmet palliative care needs might be underestimated.

## Conclusion

This study has developed empirical methods for estimating unmet palliative care needs at a population-level. We show that prevalence of unmet needs is high regardless of estimate used and projected to increase by 2050. Our study contrasts estimation methods based on unresolved symptoms and concerns and insufficient care provision for planning and evaluating of equitable care at the end of life depending on which type of metric is available at population-level.

## Data Availability

All data for the population estimates and projections are publicly available on the ONS website (population estimates: https://www.nomisweb.co.uk/datasets/mortsa; projections: https://www.ons.gov.uk/peoplepopulationandcommunity/populationandmigration/populationprojections/datasets/2014basednationalpopulationprojectionstableofcontents).

## Data Availability

All data for the population estimates and projections are publicly available on the ONS website (population estimates: https://www.nomisweb.co.uk/datasets/mortsa; projections: https://www.ons.gov.uk/peoplepopulationandcommunity/populationandmigration/populationprojections/datasets/2014basednationalpopulationprojectionstableofcontents).
Survey data may be made available upon reasonable request to Katherine Sleeman, Cicely Saunders Institute.

## Author declarations

### Data sharing statement

All data for the population-level estimates and projections are publicly available on the ONS website (2022 mortality registry data: https://www.nomisweb.co.uk/datasets/mortsa; national mortality projections:https://www.ons.gov.uk/peoplepopulationandcommunity/populationandmigration/populationprojections/datasets/2014basednationalpopulationprojectionstableofcontents).

### Contributors

Contributors TJ, AEB and FEMM accept full responsibility for the finished work and the conduct of the study, analysed the data and controlled the decision to publish. AEB, FEMM, KES, AF, TJ, JMD, LF and IJH contributed to the conception and design of the study. TJ and KES acquired and curated the Better End of Life Survey data. TJ and AEB performed the statistical analyses and drafted the manuscript. TJ, AEB, FEMM, AF, KES, JMD, IJH, LF and MD contributed to analysis and interpretation of the data and critically reviewed the manuscript for important intellectual content. TJ, AEB and FEMM are guarantors for the overall content. The corresponding author (AB) attests that all listed authors meet authorship criteria and that no others meeting the criteria have been omitted.

The authors would like to acknowledge Joel Bates for his contributions to data visualisation, and Margaret Ogden, David Chandler, and Pam Smith from our Public and Patient Involvement (PPI) group for critical input to refining estimation methods and interpretation of findings.

