## Supplement files 1-9 for "Unmet palliative care needs in England and Wales: population-based estimates and future projections (2025-2050)"

### S1. Survey sampling frame

**Supplement table 1.** ICD-10 codes for causes of death used in the sampling framework for the Better End of Life survey.

| **Cause of death** | **ICD-10 code** |
| --- | --- |
| Malignant neoplasm | C00-C97 |
| Heart disease | I00-I52, I60-I69 |
| Renal disease | N17, N18, N28 |
| Liver disease | K70-K77 |
| Respiratory disease | J06-J18, J20-J22, J40-J47, J69 |
| Neurological disease | G10, G12.2, G20, G23.1, G35, G90.3 |
| Dementia | F01, F03, G30, R54 |
| HIV/AIDS | B20-B24 |

### S2. Estimation method 1

**Supplement figure 2.** Distribution of total IPOS scores in the full sample and by nation (reference line represents a score of 34, used as a cut-off to indicate unmet palliative care needs).

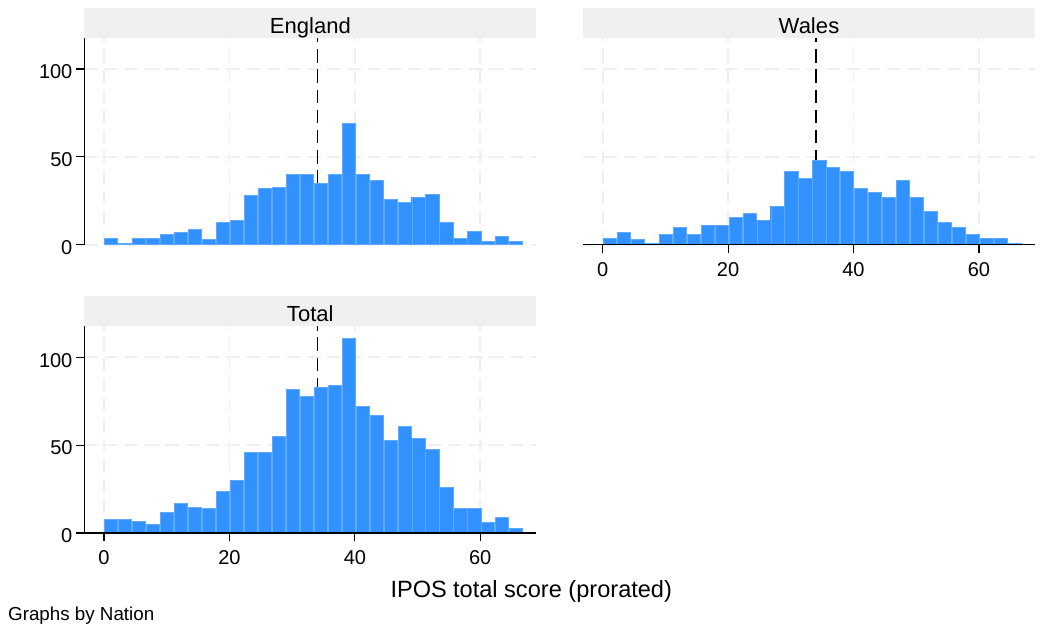

**Supplement table 2A.** Number of completed IPOS items in the survey sample (n=1194). Observations with eight or more IPOS items missing were excluded from analysis.

| **IPOS items completed** | **n** | **%** |
| --- | --- | --- |
| 17 | 836 | 70.02 |
| 16 | 119 | 9.97 |
| 15 | 55 | 4.61 |
| 14 | 36 | 3.02 |
| 13 | 30 | 2.51 |
| 12 | 17 | 1.42 |
| 11 | 18 | 1.51 |
| 10 | 25 | 2.09 |
| 9 | 16 | 1.34 |
| 8 | 6 | 0.5 |
| 7 | 13 | 1.09 |
| 6 | 2 | 0.17 |
| 5 | 1 | 0.08 |
| 3 | 1 | 0.08 |
| 2 | 3 | 0.25 |
| 1 | 4 | 0.34 |
| 0 | 12 | 1.01 |

**Sensitivity analysis**

Compared to the sample used in the main analysis, decedents in the group with less than nine completed IPOS items were more likely to have lived comfortably financially and have dementia as cause of death. It was also more common for respondents to not have been involved in informal caregiving in the last three months of their relative’s life.

**Supplement table 2B.** Descriptives for excluded sub-sample with less than nine completed IPOS items compared to sample in main analysis (nine or more completed IPOS items).

|  | **Excluded group**  **(n=42)** | **Sample in main analysis**  **(n=1152)** | **Chi-2 *p* value** |
| --- | --- | --- | --- |
| **Gender** |  |  | 0.014 |
| Male | 28 (66.7%) | 543 (47.5%) |  |
| Female | 14 (33.3%) | 601 (52.5%) |  |
| **Age** |  |  | 0.84 |
| 18-64 | 4 (9.8%) | 105 (9.2%) |  |
| 65-84 | 20 (48.8%) | 509 (44.6%) |  |
| 85 or older | 17 (41.5%) | 526 (46.1%) |  |
| **Ethnicity** |  |  | 0.32 |
| White | 41 (100.0%) | 1122 (97.7%) |  |
| Non-white | 0 (0.0%) | 27 (2.3%) |  |
| **Financial circumstances** |  |  | 0.18 |
| Living comfortably | 22 (61.1%) | 523 (46.0%) |  |
| Doing alright | 8 (22.2%) | 390 (34.3%) |  |
| Just about getting by or finding it difficult | 6 (16.7%) | 225 (19.8%) |  |
| **Nation** |  |  | 0.58 |
| England | 20 (50.0%) | 591 (51.9%) |  |
| Wales | 20 (50.0%) | 547 (48.1%) |  |
| **Cause of death** |  |  | 0.002 |
| Cancer | 3 (8.8%) | 337 (29.5%) |  |
| Heart disease | 5 (14.7%) | 167 (14.6%) |  |
| Stroke | 1 (2.9%) | 39 (3.4%) |  |
| Lung disease | 1 (2.9%) | 58 (5.1%) |  |
| Liver disease | 2 (5.9%) | 17 (1.5%) |  |
| Kidney disease | 0 (0.0%) | 8 (0.7%) |  |
| Neurological disease | 3 (8.8%) | 18 (1.6%) |  |
| Dementia | 10 (29.4%) | 167 (14.6%) |  |
| Other | 9 (26.5%) | 333 (29.1%) |  |
| **Comorbidities** |  |  | 0.019 |
| None | 22 (52.4%) | 393 (34.1%) |  |
| 1 condition | 15 (35.7%) | 377 (32.7%) |  |
| 2 conditions | 4 (9.5%) | 236 (20.5%) |  |
| 3 conditions or more | 1 (2.4%) | 146 (12.7%) |  |
| **Respondent help care for decedent in the last 3 months** |  |  | <0.001 |
| Yes | 17 (47.2%) | 877 (76.5%) |  |
| No | 19 (52.8%) | 270 (23.5%) |  |

### S3. Estimation method 2

**Supplement table 3.** Response distribution to survey question about receiving enough help from GPs in the survey sample (n=1,194).

| *Overall, do you feel your relative got as much help as needed from GPs?* | **n** | **%** |
| --- | --- | --- |
| Yes, most of the time | 482 | 40.4% |
| Sometimes | 288 | 24.1% |
| No | 213 | 17.8% |
| Did not receive care from them | 165 | 13.8% |
| Missing | 46 | 3.9% |

### S4. Unmet palliative care needs identified by estimation methods 1 and 2

**Supplement table 4.** Unmet palliative care needs identified using estimation methods 1 and 2 (excluding observations with missing data). Cell d shows unmet needs using estimation method 3 (the conservative estimate), and cells b+c+d represent unmet needs identified by estimation method 4 (the broad estimate).

|  | | | **Estimation method 2: Insufficient care provision** | | |
| --- | --- | --- | --- | --- | --- |
|  |  |  | No unmet needs | Unmet needs | Total |
| **Estimation method 1: Unresolved symptoms and concerns** | No unmet needs | *n* | 218 | 209 | 427 |
|  |  | *row %* | 51.1% | 49.0% | 100% |
|  |  | *col %* | 46.8% | 32.8%  b | 38.7% |
|  | Unmet needs | *n* | 248  a | 429 | 677 |
|  |  | *row %* | 36.6% | 63.4% | 100% |
|  |  | *col %* | 53.2%  c | 67.2%  d | 61.3% |
|  | Total | *n* | 466 | 638 | 1,104 |
|  |  | *row %* | 42.2% | 57.8% | 100% |
|  |  | *col %* | 100% | 100% | 100% |

### S5. Unmet needs by decedent characteristics (estimates 3 and 4)

| **Supplement table 5.** Frequency count and row percent of unmet palliative care needs using the conversative estimate (estimate 3) and broad estimate (estimate 4) by decedent characteristics in the Better End of Life survey sample (N=1,194). | | | | | | | | | |
| --- | --- | --- | --- | --- | --- | --- | --- | --- | --- |
|  | **(3) Conservative estimate** | | | | | **(4) Broad estimate** | | | |
|  | **No unmet needs (n=684)** | | **Unmet needs (n=437)** | | **Missing (n=73)** | **No unmet needs (n=224)** | **Unmet needs (n=897)** | | **Missing (n=73)** |
|  |  | | *n (row %)* | |  |  | *n (row %)* | |  |
| **Gender** | |  | | | |  | |  | |
| Male | 325 (47.5%) | | 205 (46.9%) | | 41 (56.2%) | 109 (48.7%) | 421 (46.9%) | | 41 (56.2%) |
| Female | 357 (52.2%) | | 226 (51.7%) | | 32 (43.8%) | 113 (50.4%) | 470 (52.4%) | | 32 (43.8%) |
| Missing | 2 (0.3%) | | 6 (1.4%) | | - | 2 (0.9%) | 6 (0.7%) | | - |
| **Age** |  | |  | |  |  |  | |  |
| 18-64 | 52 (7.6%) | | 46 (10.5%) | | 11 (15.1%) | 14 (6.2%) | 84 (9.4%) | | 11 (15.1%) |
| 65-84 | 276 (40.4%) | | 220 (50.3%) | | 33 (45.2%) | 72 (32.1%) | 424 (47.3%) | | 33 (45.2%) |
| 85 or older | 349 (51.0%) | | 166 (38.0%) | | 28 (38.4%) | 134 (59.8%) | 381 (42.5%) | | 28 (38.4%) |
| Missing | 7 (1.0%) | | 5 (1.1%) | | 1 (1.4%) | 4 (1.8%) | 8 (0.9%) | | 1 (1.4%) |
| **Ethnicity** | |  | | | |  | |  | |
| White | 668 (97.7%) | | 425 (97.3%) | | 70 (95.9%) | 222 (99.1%) | 871 (97.1%) | | 70 (95.9%) |
| Non-white | 15 (2.2%) | | 10 (2.3%) | | 2 (2.7%) | 2 (0.9%) | 23 (2.6%) | | 2 (2.7%) |
| Missing | 1 (0.1%) | | 2 (0.5%) | | 1 (1.4%) | 0 (0.0%) | 3 (0.3%) | | 1 (1.4%) |
| **Financial circumstances** | | | |  | |  | | | |
| Living comfortably | 342 (50.0%) | | 167 (38.2%) | | 36 (49.3%) | 124 (55.4%) | 385 (42.9%) | | 36 (49.3%) |
| Doing alright | 220 (32.2%) | | 159 (36.4%) | | 19 (26.0%) | 69 (30.8%) | 310 (34.6%) | | 19 (26.0%) |
| Just about getting by or finding it difficult | 115 (16.8%) | | 104 (23.8%) | | 12 (16.4%) | 30 (13.4%) | 189 (21.1%) | | 12 (16.4%) |
| Missing | 7 (1.0%) | | 7 (1.6%) | | 6 (8.2%) | 1 (0.4%) | 13 (1.4%) | | 6 (8.2%) |
| **Nation** | |  | | | |  | |  | |
| England | 340 (49.7%) | | 243 (55.6%) | | 36 (49.3%) | 117 (52.2%) | 466 (52.0%) | | 36 (49.3%) |
| Wales | 344 (50.3%) | | 194 (44.4%) | | 37 (50.7%) | 107 (47.8%) | 431 (48.0%) | | 37 (50.7%) |
| **Cause of death** | |  | | | |  | |  | |
| Cancer | 188 (27.5%) | | 141 (32.3%) | | 11 (15.1%) | 46 (20.5%) | 283 (31.5%) | | 11 (15.1%) |
| Heart disease | 121 (17.7%) | | 46 (10.5%) | | 5 (6.9%) | 37 (16.5%) | 130 (14.5%) | | 5 (6.9%) |
| Stroke | 28 (4.1%) | | 10 (2.3%) | | 2 (2.7%) | 6 (2.7%) | 32 (3.6%) | | 2 (2.7%) |
| Lung disease | 31 (4.5%) | | 25 (5.7%) | | 3 (4.1%) | 17 (7.6%) | 39 (4.3%) | | 3 (4.1%) |
| Liver disease | 5 (0.7%) | | 10 (2.3%) | | 4 (5.5%) | 0 (0.0%) | 15 (1.7%) | | 4 (5.5%) |
| Kidney disease | 4 (0.6%) | | 3 (0.7%) | | 1 (1.4%) | 3 (1.3%) | 4 (0.4%) | | 1 (1.4%) |
| Neurological disease | 14 (2.0%) | | 4 (0.9%) | | 3 (4.1%) | 7 (3.1%) | 11 (1.2%) | | 3 (4.1%) |
| Dementia | 106 (15.5%) | | 52 (11.9%) | | 19 (26.0%) | 42 (18.8%) | 116 (12.9%) | | 19 (26.0%) |
| Other | 185 (27.0%) | | 141 (32.3%) | | 16 (21.9%) | 65 (29.0%) | 261 (29.1%) | | 16 (21.9%) |
| Missing | 2 (0.3%) | | 5 (1.1%) | | 9 (12.3%) | 1 (0.4%) | 6 (0.7%) | | 9 (12.3%) |
| **Comorbidities** | |  | | | |  | |  | |
| None | 252 (36.8%) | | 124 (28.4%) | | 39 (53.4%) | 89 (39.7%) | 287 (32.0%) | | 39 (53.4%) |
| 1 condition | 223 (32.6%) | | 144 (33.0%) | | 25 (34.3%) | 73 (32.6%) | 294 (32.8%) | | 25 (34.3%) |
| 2 conditions | 129 (18.9%) | | 103 (23.6%) | | 8 (11.0%) | 40 (17.9%) | 192 (21.4%) | | 8 (11.0%) |
| 3 conditions or more | 80 (11.7%) | | 66 (15.1%) | | 1 (1.4%) | 22 (9.8%) | 124 (13.8%) | | 1 (1.4%) |

### S6. Age-, gender- and nation-specific sample estimates

**Supplement table 6.** Age-, gender- and nation-specific sample estimates of unmet palliative care needs among decedents in the Better End of Life survey sample (with 95% confidence intervals).

| **England** | | | | | | | |
| --- | --- | --- | --- | --- | --- | --- | --- |
| **Estimate** |  | **Male** | | | **Female** | | |
|  | **Age** | *n* | *%* | *95%CI (LCI*–*UCI)* | *n* | *%* | *95%CI (LCI*–*UCI)* |
| **1. Unresolved symptoms and concerns** | **18-64** | 26 | 65.0 | 50.2–79.8 | 16 | 76.2 | 58.0–94.4 |
|  | **65-84** | 96 | 71.1 | 63.5–78.8 | 76 | 61.8 | 53.2–70.4 |
|  | ≥**85** | 48 | 44.9 | 35.4–54.3 | 93 | 56.4 | 48.8–63.9 |
| **2. Insufficient care provision** | **18-64** | 23 | 60.5 | 45.0–76.1 | 10 | 50.0 | 28.1–71.9 |
|  | **65-84** | 80 | 58.8 | 50.6–67.1 | 84 | 67.2 | 59.0–75.4 |
|  | ≥**85** | 64 | 58.7 | 49.5–68.0 | 96 | 59.6 | 52.0–67.2 |
| **3. Conservative estimate (1 and 2 combined)** | **18-64** | 16 | 44.4 | 28.2–60.7 | 10 | 50.0 | 28.1–71.9 |
|  | **65-84** | 61 | 46.6 | 38.0–55.1 | 51 | 41.8 | 33.1–50.6 |
|  | ≥**85** | 36 | 33.6 | 24.7–42.6 | 63 | 39.6 | 32.0–47.2 |
| **4. Broad estimate (either 1 or 2)** | **18-64** | 31 | 86.1 | 74.8–97.4 | 15 | 75.0 | 56.0–94.0 |
|  | **65-84** | 109 | 83.2 | 76.8–89.6 | 107 | 87.7 | 81.9–93.5 |
|  | ≥**85** | 75 | 70.1 | 61.4–78.8 | 122 | 76.7 | 70.2–83.3 |

| **Wales** | | | | | | | |
| --- | --- | --- | --- | --- | --- | --- | --- |
| **Estimate** |  | **Male** | | | **Female** | | |
|  | **Age** | *n* | *%* | *95%CI (LCI*–*UCI)* | *n* | *%* | *95%CI (LCI*–*UCI)* |
| **1. Unresolved symptoms and concerns** | **18-64** | 14 | 58.3 | 38.6–78.1 | 14 | 73.7 | 53.9–93.5 |
|  | **65-84** | 96 | 70.6 | 62.9–78.2 | 81 | 70.4 | 62.1–78.8 |
|  | ≥**85** | 53 | 54.6 | 44.7–64.5 | 83 | 54.2 | 46.4–62.1 |
| **2. Insufficient care provision** | **18-64** | 18 | 72.0 | 54.4–89.6 | 12 | 66.7 | 44.9–88.4 |
|  | **65-84** | 78 | 57.4 | 49.0–65.7 | 71 | 63.4 | 54.5–72.3 |
|  | ≥**85** | 46 | 46.9 | 37.1–56.8 | 73 | 48.0 | 40.1–56.0 |
| **3. Conservative estimate (1 and 2 combined)** | **18-64** | 10 | 43.5 | 23.2–63.7 | 9 | 50.0 | 26.9–73.1 |
|  | **65-84** | 53 | 39.8 | 31.5–48.2 | 55 | 50.0 | 40.7–59.3 |
|  | ≥**85** | 29 | 30.2 | 21.0–39.4 | 36 | 24.2 | 17.3–31.0 |
| **4. Broad estimate (either 1 or 2)** | **18-64** | 20 | 87.0 | 73.2–100.7 | 17 | 94.4 | 83.9–105.0 |
|  | **65-84** | 116 | 87.2 | 81.5–92.9 | 92 | 83.6 | 76.7–90.5 |
|  | ≥**85** | 67 | 69.8 | 60.6–79.0 | 115 | 77.2 | 70.4–83.9 |

Notes: Sample estimates are based on denominators excluding observations with missing data.

### S7. Regression results

**Supplement table S7.** Results from the modified Poisson regression models, showing main effects (unadjusted and adjusted prevalence ratios (PR) with 95% confidence intervals) for associations between decedent characteristics and risk of unmet palliative care needs using estimation methods 1 and 2.

|  | **Method 1: Unresolved symptoms and concerns** | | **Method 2: Insufficient care provision** | | |
| --- | --- | --- | --- | --- | --- |
|  | **Unadjusted PR** [95% CI] | **Adjusted PR** [95% CI] | | **Unadjusted PR** [95% CI] | **Adjusted PR** [95% CI] |
| **Gender** |  |  | |  |  |
| Male (reference group) | **-** | **-** | | **-** | **-** |
| Female | 0.99 | 0.99 | | 1.03 | 1.06 |
|  | [0.90,1.08] | [0.91,1.09] | | [0.93,1.14] | [0.96,1.18] |
| **Age** |  |  | |  |  |
| 18-64 | 1.27** | 1.15 | | 1.17 | 1.15 |
|  | [1.09,1.48] | [0.98,1.35] | | [0.99,1.39] | [0.96,1.39] |
| 65-84 | 1.29*** | 1.20*** | | 1.15* | 1.17** |
|  | [1.17,1.43] | [1.09,1.33] | | [1.03,1.27] | [1.04,1.30] |
| ≥85 (reference group) | - | - | | - | - |
| **Ethnicity** |  |  | |  |  |
| White (reference group) | - | - | | - | - |
| Non-white | 1.22 | 1.11 | | 1.04 | 0.91 |
|  | [0.97,1.53] | [0.88,1.41] | | [0.75,1.43] | [0.64,1.28] |
| **Financial circumstances** |  |  | |  |  |
| Living comfortably (reference group) | - | - | | - | - |
| Doing alright | 1.21*** | 1.19** | | 1.07 | 1.05 |
|  | [1.09,1.35] | [1.07,1.33] | | [0.95,1.20] | [0.94,1.18] |
| Just about getting by or finding it difficult | 1.37*** | 1.29*** | | 1.13 | 1.08 |
|  | [1.22,1.53] | [1.15,1.44] | | [0.99,1.28] | [0.95,1.24] |
| **Nation** |  |  | |  |  |
| England (reference group) | - | - | | - | - |
| Wales | 1.02 | 1.054 | | 0.08* | 0.90 |
|  | [0.93,1.12] | [0.96,1.15] | | [0.81,0.99] | [0.82,1.00] |
| **Cause of death** |  |  | |  |  |
| Cancer | 1.21*** | 1.22*** | | 0.94 | 0.90 |
|  | [1.09,1.35] | [1.09,1.36] | | [0.83,1.05] | [0.79,1.02] |
| Heart disease | 0.87 | 0.87 | | 0.88 | 0.89 |
|  | [0.73,1.03] | [0.74,1.03] | | [0.75,1.03] | [0.76,1.04] |
| Dementia | 0.99 | 1.03 | | 0.80** | 0.81* |
|  | [0.85,1.15] | [0.89,1.21] | | [0.67,0.94] | [0.68,0.96] |
| Lung disease | 0.94 | 0.94 | | 0.96 | 0.98 |
|  | [0.73,1.20] | [0.74,1.19] | | [0.77,1.20] | [0.78,1.22] |
| Other conditions (reference group) | - | - | | - | - |
| **Comorbidities** |  |  | |  |  |
| None (reference group) | - | - | | - | - |
| 1 condition | 1.17* | 1.18** | | 1.04 | 1.05 |
|  | [1.04,1.33] | [1.04,1.33] | | [0.92,1.17] | [0.92,1.18] |
| 2 conditions | 1.29*** | 1.30*** | | 1.06 | 1.04 |
|  | [1.13,1.47] | [1.14,1.49] | | [0.92,1.21] | [0.90,1.19] |
| 3 conditions or more | 1.44*** | 1.44*** | | 0.96 | 0.90 |
|  | [1.26,1.65] | [1.26,1.66] | | [0.80,1.13] | [0.75,1.07] |
| **Constant** |  | 0.39 | |  | 0.56 |
|  |  | [0.33, 0.46] | |  | [0.48, 0.65] |

* *p* < 0.05, ** *p* < 0.01, *** *p* < 0.001

### S8. Age-, gender-, and nation-specific population estimates

**Supplement table S8.** Age-, gender- and nation-specific population-level estimates of unmet palliative care needs (count and percentage of all deaths, with lower and upper bounds based on lower and upper 95% confidence for each sample estimate).

| **England** | | | | | |
| --- | --- | --- | --- | --- | --- |
| **Estimate** | **Age groups** | **Male** | | **Female** | |
|  |  | Estimated *n (%)* | Lower– Upper estimate *(%)* | Estimated *n (%)* | Lower–Upper estimate *(%)* |
| **1. Unresolved symptoms and concerns** | **20-64** | 20,592 (42.6) | 15,909–25,275 (32.9–52.3) | 17,000 (54.4) | 12,935–21,064 (41.4–67.4) |
|  | **65-84** | 75,020 (55.8) | 66,954–83,086 (49.8–61.8) | 52,534 (48.5) | 45,233–59,835 (41.7–55.2) |
|  | ≥**85** | 29,667 (33.5) | 23,435–35,899 (26.5–40.6) | 53,180 (42.4) | 46,040–60,320 (36.7–48.1) |
| **2. Insufficient care provision** | **20-64** | 19,175 (39.7) | 14,251–24,098 (29.5–52.3) | 11,156 (35.7) | 6,267–16,045 (20.1–51.4) |
|  | **65-84** | 62,057 (46.2) | 53,331–70,783 (39.7–61.8) | 57,135 (52.7) | 50,137–64,132 (46.3–59.2) |
|  | ≥**85** | 38,830 (43.9) | 32,717–44,942 (37.0–50.8) | 56,260 (44.8) | 49,109–63,410 (39.1–50.5) |
| **3. Conservative estimate (1 and 2 combined)** | **20-64** | 14,080 (29.1) | 8,938–19,222 (18.5–31.8) | 11,156 (35.7) | 6,267–16,045 (20.1–51.4) |
|  | **65-84** | 49,125 (36.6) | 40,113–58,136 (29.8–43.3) | 35,542 (32.8) | 28,100–42,984 (25.9–39.7) |
|  | ≥**85** | 22,250 (25.1) | 16,329–28,171 (18.5–31.8) | 37,385 (29.8) | 30,211–44,558 (24.1–35.5) |
| **4. Broad estimate (either 1 or 2)** | **20-64** | 27,280 (56.4) | 23,701–30,859 (49.0–63.8) | 16,734 (53.6) | 12,500–20,968 (64.2–73.4) |
|  | **65-84** | 87,780 (65.3) | 81,027–94,533 (60.3–70.3) | 74,568 (68.8) | 69,614–79,523 (64.2–73.4) |
|  | ≥**85** | 46,354 (52.4) | 40,617–52,091 (45.9–58.9) | 72,396 (57.7) | 66,199–78,593 (52.7–62.6) |

| **Wales** | | | | | |
| --- | --- | --- | --- | --- | --- |
| **Estimate** | **Age groups** | **Male** | | **Female** | |
|  |  | *Estimated n (%)* | Lower–Upper estimate *(%)* | *Estimated n (%)* | Lower–Upper estimate *(%)* |
| **1. Unresolved symptoms and concerns** | **20-64** | 1,137 (37.6) | 752–1,521  (24.9–50.3) | 1,106  (52.9) | 809–1,403  (38.7–67.2) |
|  | **65-84** | 5,178 (55.8) | 4,616–5,739  (49.8–61.9) | 4,255  (55.2) | 3,751–4,759  (48.6–61.7) |
|  | ≥**85** | 2,235 (41.1) | 1,830–2,640  (33.6–48.5) | 3,298  (41.4) | 2,818–3,778  (35.3–47.4) |
| **2. Insufficient care provision** | **20-64** | 1,403 (46.4) | 1,060–1,746  (35.0–57.7) | 1,001  (47.9) | 674–1,328  (32.3–63.5) |
|  | **65-84** | 4,207 (45.4) | 3,597–4,817  (38.8–52.0) | 3,830  (49.7) | 3,291–4,369  (42.7–56.6) |
|  | ≥**85** | 1,920 (35.3) | 1,516–2,324  (27.9–42.7) | 2,920  (36.6) | 2,437–3,403  (30.6–42.7) |
| **3. Conservative estimate (1 and 2 combined)** | **20-64** | 847  (28.0) | 453–1,242  (15.0–41.1) | 751  (35.9) | 404–1,097  (19.3–52.5) |
|  | **65-84** | 2,923 (31.5) | 2,313–3,533  (24.9–38.1) | 3,021  (39.2) | 2,456–3,585  (31.8–46.5) |
|  | ≥**85** | 1,236 (22.7) | 860–1,611  (15.8–29.6) | 1,469  (18.4) | 1,051–1,887  (13.2–23.7) |
| **4. Broad estimate (either 1 or 2)** | **20-64** | 1,695 (56.0) | 1,427–1,963  (47.2–64.9) | 1,418  (67.9) | 1,259–1,576  (60.3–75.5) |
|  | **65-84** | 6,397 (69.0) | 5,981–6,814  (64.5–73.5) | 5,052  (65.6) | 4,635–5,470  (60.1–70.9) |
|  | ≥**85** | 2,854 (52.5) | 2,479–3,230  (45.6–59.4) | 4,693  (58.9) | 4,283–5,102  (53.7–64.0) |

### S9. Projections of unmet palliative care needs in absolute numbers

**Supplement table S9.** Projected numbers of adult decedents with unmet palliative care needs in England and Wales by 2025 and 2050, across estimates.

|  |  | **England** | | | | **Wales** | | | |
| --- | --- | --- | --- | --- | --- | --- | --- | --- | --- |
| **Estimate** | **Age groups** | **Female** | | **Male** | | **Female** | | **Male** | |
|  |  | **2025**  **n** | **2050**  **n** | **2025**  **n** | **2050**  **n** | **2025**  **n** | **2050**  **n** | **2025**  **n** | **2050**  **n** |
| **1. Unresolved symptoms and concerns** | **20-64** | 17,875 | 15,504 | 21,595 | 18,185 | 1,149 | 980 | 1,187 | 985 |
|  | **65-84** | 54,573 | 55,280 | 78,495 | 80,604 | 4,360 | 4,178 | 5,398 | 5,208 |
|  | ≥**85** | 51,899 | 85,060 | 29,358 | 53,029 | 3,091 | 5,109 | 2,188 | 4,061 |
| **2. Insufficient care provision** | **20-64** | 11,730 | 10,174 | 20,109 | 16,933 | 1,039 | 887 | 1,465 | 1,216 |
|  | **65-84** | 59,353 | 60,121 | 64,932 | 66,676 | 3,924 | 3,760 | 4,386 | 4,232 |
|  | ≥**85** | 54,904 | 89,985 | 38,425 | 69,408 | 2,737 | 4,523 | 1,880 | 3,489 |
| **3. Conservative estimate (1 and 2 combined)** | **20-64** | 11,730 | 10,174 | 14,766 | 12,434 | 780 | 665 | 884 | 736 |
|  | **65-84** | 36,922 | 37,400 | 51,400 | 52,781 | 3,095 | 2,966 | 3,047 | 2,988 |
|  | ≥**85** | 36,484 | 59,796 | 22,018 | 39,772 | 1,377 | 2,275 | 1,210 | 2,192 |
| **4. Broad estimate (either 1 or 2)** | **20-64** | 17,596 | 15,262 | 28,609 | 24,091 | 1,473 | 1,256 | 1,769 | 1,469 |
|  | **65-84** | 77,463 | 78,466 | 91,846 | 94,313 | 5,177 | 4,961 | 6,669 | 6,435 |
|  | ≥**85** | 70,652 | 115,795 | 45,871 | 82,858 | 4,398 | 7,269 | 2,795 | 5,187 |
